# Racial and Ethnic Disparities in SARS-CoV-2 Pandemic: Analysis of a COVID-19 Observational Registry for a Diverse U.S. Metropolitan Population

**DOI:** 10.1101/2020.04.24.20073148

**Authors:** Farhaan S. Vahidy, Juan Carlos Nicolas, Jennifer R. Meeks, Osman Khan, Stephen L. Jones, Faisal N. Masud, H. Dirk Sostman, Robert A. Phillips, Julia D. Andrieni, Bita Kash, Khurram Nasir

**Author notes:** **Corresponding Author** Farhaan S. Vahidy, PhD MBBS MPH, Josie Roberts Building, Houston Methodist, 7550 Greenbriar Drive, Houston TX 77030.

## Abstract

**Introduction:** Data on race and ethnic susceptibility to SARS-CoV-2 infection are limited. We analyzed socio-demographic factors associated with higher likelihood of SARS-CoV-2 infection and explore mediating pathways for race disparities in the SARS-CoV-2 pandemic.

**Methods:** Cross sectional analysis of COVID-19 Surveillance and Outcomes Registry (CURATOR), which captures data for a large healthcare system comprising of one central tertiary care, seven large community hospitals, and an expansive ambulatory / emergency care network in the Greater Houston area. Nasopharyngeal samples for individuals inclusive of all ages, races, ethnicities and sex were tested for SARS-CoV-2. We analyzed, socio-demographic (age, sex, race, ethnicity, household income, residence population density) and comorbidity (hypertension, diabetes, obesity, cardiac disease) factors. Multivariable logistic regression models were fitted to provide adjusted Odds Ratios (aOR), 95% confidence intervals (CI) for likelihood of positive SARS-CoV-2 test. Structural Equation Modeling (SEM) framework was utilized to explore three mediation pathways (low income, high population density, high comorbidity burden) for association between African American race and SARS-CoV-2 infection.

**Results:** Among 4,513 tested individuals, 754 (16.7%) tested positive. Overall mean (SD) age was 50.6 (18.9) years, 62% females and 26% were African American. African American race was associated with lower socio-economic status, higher comorbidity burden, and population density residence. In the fully adjusted model, African American race (vs. White; aOR, CI: 1.84, 1.49-2.27) and Hispanic ethnicity (vs. non-Hispanic; aOR, CI: 1.70, 1.35-2.14) had a higher likelihood of infection. Older individuals and males were also at a higher risk of SARS-CoV-2 infection. The SEM framework demonstrated a statistically significant (p = 0.008) indirect effect of African American race on SARS-CoV-2 infection mediated via a pathway that included residence in densely populated zip code.

**Conclusions:** There is strong evidence of race and ethnic disparities in the SARS-CoV-2 pandemic potentially mediated through unique social determinants of health.

Strengths and limitations of this study
- One of the first studies to systematically evaluate race and ethnic disparities in susceptibility to SARS-CoV-2 infection, while accounting for multiple sociodemographic characteristics and comorbidities
- Study population represents a large and diverse metropolitan of the U.S. with data from one of the largest healthcare providers across the greater metropolitan area
- Study evaluates potential mediation pathways for race disparities and demonstrates that residence in areas with high population density may mediate race disparities in susceptibility to SARS-CoV-2 infection
- Single center study with limited information about true burden of comorbidity and lifestyle factors

## Introduction

The Coronavirus (COVID-19) disease caused by infection with the Severe Acute Respiratory Syndrome Coronavirus 2 (SARS-CoV-2) virus is a pandemic that has thus far resulted in over 2 million cases across 170 countries in under 4 months. At the time of this reporting, the U.S. has approximately 30% of total global cases, and has surpassed all countries in terms of absolute number of cases and fatalities.^1,2^ Experts project these numbers to continue rising as widespread testing is instituted. The geographic distribution of cases across the U.S. suggests that the major pandemic burden has hit metropolitan areas such as New York; however, cases of COVID-19 have now been reported across all 50 states, the District of Columbia, Guam, Puerto Rico, the Northern Mariana Islands, and the U.S. Virgin Islands.^3^ As of April 18, the state of Texas had 18,260 reported cases of COVID-19, with approximately one-third in the Greater Houston area.^4^ The greater Houston area is home to approximately 7 million individuals, is the fourth-largest metropolitan area by population in the U.S. and is considered one of the nation’s most diverse regions.

Initial reports from China and Europe indicate that specific individuals such as the elderly; males; and people with comorbidities including hypertension, diabetes, obesity, coronary artery disease and heart failure have poor COVID-19 outcomes.^5-8^ As the pandemic spread over the continental U.S. during the last two months, patterns of high-risk phenotypes have started to emerge, and reports of poor outcomes (particularly high case fatality) among racial minorities have surfaced in the media.^9-11^ Though it is important to understand the determinants of poor outcomes among COVID-19 patients, it is equally imperative, from a public health perspective, to systematically examine the likelihood of SARS-CoV-2 infection across large diverse communities in the U.S. More specifically, data on potential higher likelihood of SARS-CoV-2 infection among racial and ethnic minorities across diverse U.S. metropolitan areas outside of New York are limited. Furthermore, the mediators of SARS-CoV-2 infection among racial and ethnic minorities have not been described.

We explored socio-demographic characteristics such as age, sex, race, ethnicity, median household zip code income, population density of residents’ zip codes, and health insurance status associated with positive SARS-CoV-2 testing in an urban and diverse population served by one of the leading healthcare systems of the greater Houston area. We further examined the association between pre-existing comorbidities and higher likelihood of SARS-CoV-2 infection in our study population. We hypothesized that older age, non-white race and ethnic minority status will be associated with significantly higher likelihood of SARS-CoV-2 infection, and factors such as low socio-economic status, residence in high population density areas (proxy for potential difficulties in social distancing) and higher comorbidity burden will mediate the effect of race on SARS-CoV-2 infection.

## Methods

We analyzed data being contemporaneously collected since March 5, 2020 as a part of the COVID-19 Surveillance and Outcomes Registry (CURATOR) at the Houston Methodist Hospital system (HM). The Houston Methodist CURATOR has been approved by the HM Institutional Review Board (IRB) as an observational quality of care registry for all suspected and confirmed COVID-19 patients. CURATOR is populated from multiple data sources across the HM system such as electronic medical records, electronic databanks for laboratory and pharmacy, and electronic interactive patient interface tools. The HM system comprises a flagship tertiary care hospital in the Texas Medical Center, seven large community hospitals, a continuing care hospital, and multiple emergency centers and clinics throughout the Greater Houston area. Data from various sources are curated into a harmonized format, assessed for quality and integrity, and stored on a secure institutional HIPAA-compliant server.

We flagged all individuals who were tested for the SARS-CoV-2 using the real time Reverse Transcriptase (RT) Polymerized Chain Reaction (PCR) diagnostic panels. These assays were verified for quantitative detection of novel SARS-CoV-2 isolated and purified from nasopharyngeal swab specimens obtained from individuals and immersed in universal transport medium. Testing was carried out for symptomatic individuals or for individuals who had a self-reported history of exposure to a COVID-19 case including recent travel to other countries with high infection rates or hotspots within the U.S. Socio-demographic characteristics including age, sex, race, ethnicity, and payer-status (insurance type) were obtained from the HM CURATOR for analyses. We utilized the U.S. Census Bureau’s American Community Survey (ACS) 5-year data (2014-2018) to determine median household income by individual zip code tabulation areas (ZCTA).^12^ The median ZCTA household income was inflation-adjusted to 2018 USD. We also utilized the same data source to obtain population estimates by ZCTA, and calculated ZCTA level population density (population per mile square) by standardizing it for area measurements of ZCTA. For the purpose of population density determination, land area estimates were obtained from the Census Bureau’s U.S. Gazetteer Files 2010.^13^ In the absence of granular and precise social distancing data, we have utilized population density as a proxy for potential difficulties in social distancing among crowded communities.

We provide descriptive summary data as means (standard deviations) and proportions. We fit univariable and multivariable logistic regression models to assess unadjusted and adjusted association between socio-demographic characteristics and likelihood of being tested positive for the SARS-CoV-2. We determined *a priori* to include all variables (age, sex, race, ethnicity, zip code household income, insurance type, zip population density and comorbidities) in our final multivariable model. We assessed the model fit utilizing the Hosmer and Lemeshow goodness of fit test and crude and adjusted odds ratios (OR and aOR), and 95% confidence intervals (CI) are reported. Age, income and population density variables were categorized to improve model fit. Post-estimation marginal probabilities of SARS-CoV-2 infection were determined from the fully adjusted model for major covariates (race, ethnicity and age). A comorbidity burden score was calculated by assigning one point each for presence of hypertension, diabetes, obesity or a combination of Coronary Artery Disease / Myocardial Infarction / Congestive Heart Failure (CAD / MI / CHF). We explored the mediation influence of comorbidity burden, socio-economic status (median income), and lack of social distancing (population density) on the relationship between African American race and high likelihood of SARS-CoV-2 infection using the Generalized Structural Equation Modeling (GSEM) framework. The GSEM framework was set up to provide estimates of direct and indirect effect of African American race on SARS-CoV-2 infectivity. Statistically significant (p < 0.05) indirect effects represent full or partial mediation by a tested covariate. We included all individuals tested for SARS-CoV-2 across our healthcare system and did not perform formal sample size calculations.

Patient and public involvement: There was no direct patient involvement in the design and conduct of this study.

## Results

From the HM CURATOR, during an approximate 5-week (37-day) time period, we identified a total of 4,513 presumed cases tested for SARS-CoV-2, among whom 754 (16.7%, 95% CI: 15.6 - 17.8) tested positive. Figure 1 represents temporal course of total, positive, and negative SARS-CoV-2 tests across the 37-day timeline in our hospital system.

**Figure 1:**
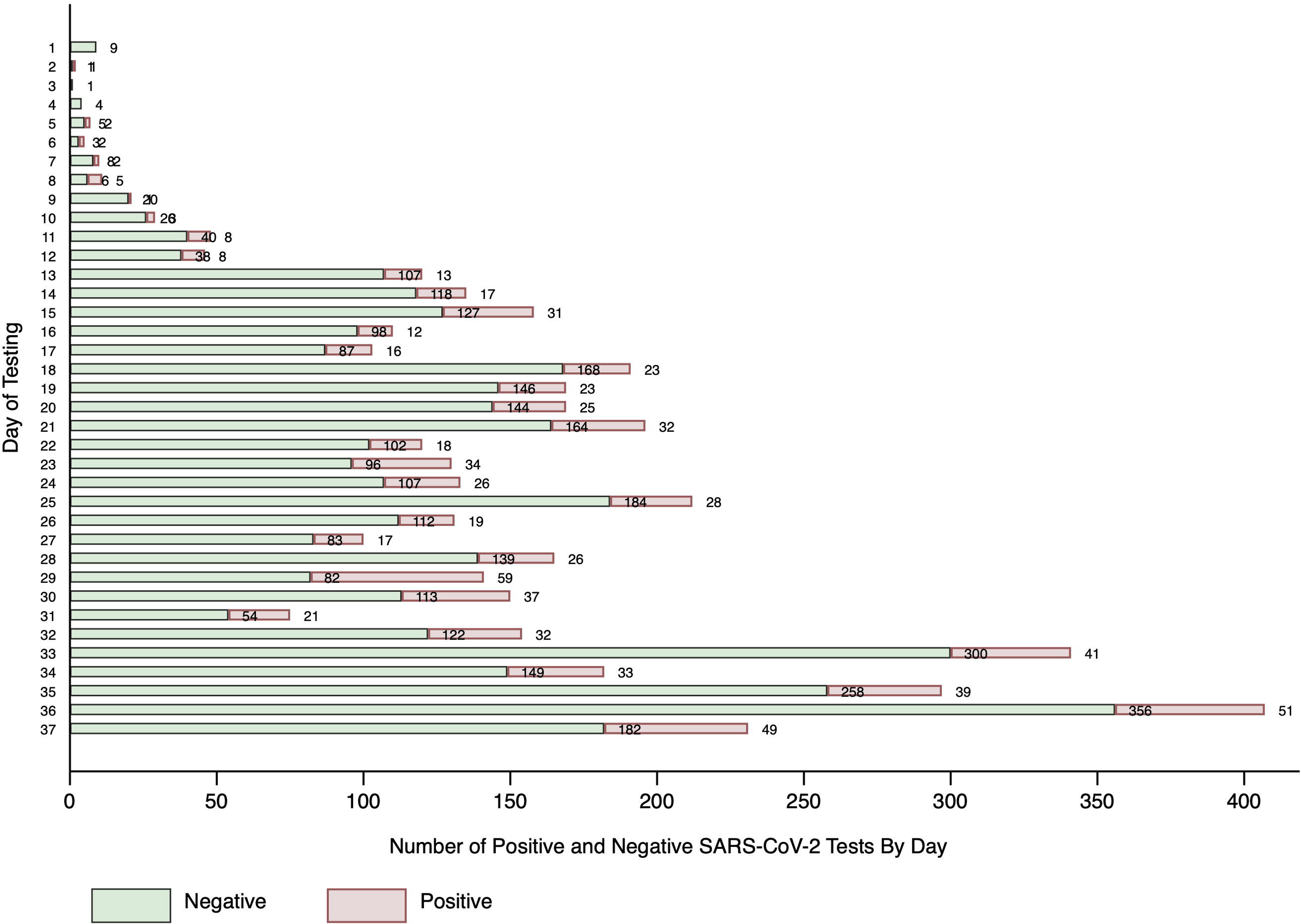
Schematic representation of the temporal sequence for total, positive and negative numbers of SARS-CoV-2 tests in the Houston Methodist CURATOR

### Socio-Demographic and Comorbidity Characteristics of the Study Population

Overall, the mean (SD) age of the study population was 50.6 (18.9) years; 62% were female and 58% were Caucasian. The overall median (IQR) household income was USD $70,324 ($53,116-$97,747), and 39.8% of the study population had private or employer-based insurance. In our univariate analysis, African American race (vs. White; OR, CI: 1.52, 1.28–1.82), Hispanic (vs. non-Hispanic; OR, CI: 1.26, 1.04-1.54), and males (vs. females; OR, CI: 1.30, 1.11-1.51), were associated with significantly higher likelihood of testing positive for SARS-CoV-2. Furthermore, among the SARS-CoV-2 positive patients, 44% were in the age category of 51-75 years, and 11% were greater than 75 years. These proportions were significantly higher than the reference group (up to 35 years; OR, CI for 51-75 years vs. up to 35 years: 1.76, 1.42-2.18 and for >75 years vs. up to 35 years: 1.35, 1.01-1.79). Furthermore, individuals in higher pentiles of socio-economic status had significantly lower likelihood; whereas, those residing in higher population density ZCTAs had higher likelihood of SARS-CoV-2 infection. Among comorbidities, a significantly greater proportion of diabetic individuals had SARS-CoV-2 positive results (OR, CI: 1.40, 0.17 - 1.68). The socio-demographic characteristics and comorbidity profiles for the overall and SARS-CoV-2 positive and negative patients are summarized in Table 1.

**Table 1:** Summary measures and univariable association of socio-demographic characteristics with SARS-CoV-2 infection from HM CURATOR

| Characteristics | Overall<br>(n = 4,513) | SARS-CoV-2<br>Negative<br>(n = 3,759) | SARS-CoV-2<br>Positive<br>(n = 754) | OR (95% CI) |
| --- | --- | --- | --- | --- |
| Age, mean (SD) | 50.6 (18.9) | 50.1 (19.1) | 53.2 (17.8) | 1.01 (1.00–1.01) |
| Age Categories (%) |  |  |  |  |
| Up to 35 years | 24.8 | 26.0 | 19.0 | Reference Category |
| 36–50 years | 27.4 | 27.8 | 25.3 | 1.25 (0.99–1.58) |
| 51–75 years | 35.9 | 34.3 | 44.0 | 1.76 (1.42–2.18) |
| >75 years | 11.9 | 11.9 | 11.7 | 1.35 (1.01–1.79) |
| Females (%) | 62.1 | 63.2 | 56.9 | 0.77 (0.66–0.90) |
| Race (%) |  |  |  |  |
| White | 57.7 | 58.8 | 51.7 | Reference Category |
| African American | 25.7 | 24.3 | 32.6 | 1.52 (1.28–1.82) |
| Asian | 9.4 | 9.2 | 10.3 | 1.27 (0.97–1.67) |
| More Than One Race /<br>Other / Unknown | 7.3 | 7.7 | 5.2 | 0.76 (0.54–1.08) |
| Hispanic (%)* | 18.7 | 18.1 | 21.8 | 1.26 (1.04–1.54) |
| Median Zip Household<br>Income (IQR)† | 70,324<br>(53,116–97,747) | 70,658<br>(53,313–97,747) | 66,983<br>(50,665–95,835) | -3675‡<br>(-6667.01, -682.95) |
| Median Zip Household Income Pentiles (%) |  |  |  |  |
| I: 24,993–50,462 | 19.8 | 19.1 | 23.6 | Reference Category |
| II: 50,465–65,339 | 18.5 | 18.4 | 19.4 | 0.85 (0.67–1.09) |
| III: 65,742–78,487 | 21.6 | 21.9 | 20.3 | 0.75 (0.59–0.95) |
| IV: 78,06–102,583 | 19.7 | 20.3 | 16.8 | 0.67 (0.52–0.86) |
| V: 103,48–230,750 | 20.4 | 20.4 | 19.9 | 0.79 (0.62–1.00) |
| Median (IQR) Population<br>Density | 2991.6<br>(1538.9 – 4299.7) | 2883.9<br>(1504 – 4261) | 3320.3<br>(2123.6 – 4665.6) | 436.4‡<br>(265.7 – 607.1) |
| Median Population Density Pentiles (%) |  |  |  |  |
| I: 1.5 – 1370.7 | 19.9 | 20.8 | 15.8 | Reference Category |
| II: 1393.7 – 2524.0 | 19.5 | 19.8 | 18.0 | 1.19 (0.91 – 1.56) |
| III: 2603.0 – 3486.5 | 20.4 | 20.1 | 22.0 | 1.44 (1.11 – 1.86) |
| IV: 3513.1 – 4789.7 | 19.8 | 19.7 | 20.4 | 1.36 (1.05 – 1.76) |
| V: 4801.1 – 61610.2 | 20.4 | 19.8 | 23.7 | 1.57 (1.22 – 2.03) |
| Insurance Status (%) |  |  |  |  |
| Medicare | 28.8 | 28.5 | 30.1 | Reference Category |
| Medicaid | 3.6 | 3.8 | 2.5 | 0.63 (0.38–1.04) |
| Pvt / Employer based | 39.8 | 39.7 | 40.2 | 0.96 (0.79–1.16) |
| HC Exchange | 2.4 | 2.2 | 3.6 | 1.57 (0.99–2.49) |
| Self-Pay | 24.3 | 24.5 | 23.2 | 0.90 (0.72–1.11) |
| VA | 1.2 | 1.3 | 0.4 | 0.28 (0.09–0.92) |
| Hypertension | 42.3 | 41.9 | 44.4 | 1.11 (0.95 – 1.30) |
| Diabetes | 21.7 | 20.7 | 26.8 | 1.40 (1.17 – 1.68) |
| Obesity | 8.0 | 8.1 | 7.7 | 0.92 (0.68 – 1.23) |

| **Characteristics** | **Overall**<br>**(n = 4,513)** | **SARS-CoV-2**<br>**Negative**<br>**(n = 3,759)** | **SARS-CoV-2**<br>**Positive**<br>**(n = 754)** | **OR (95% CI)** |
| --- | --- | --- | --- | --- |
| CAD / MI / CHF | 16.5 | 16.8 | 15.0 | 0.88 (0.70 – 1.09) |

\*Missing, Unknown, Declined n = 154 (3.4%).
†2018 inflation adjusted USD. Missing n = 76 (1.7%)
‡ Difference in median and 95% CI of difference obtained via quantile regression

### Socio-demographic and comorbidity characteristics associated with African American Race

In order to understand the association between African American race and other sociodemographic factors, we compared age, sex, median income, population density, and comorbidity profile between African American and non-African American race. Although African Americans had higher proportion of younger individuals and greater proportion of females. A significantly higher proportion of African Americans had lower socio-economic status, resided in ZCTAs with higher population density, and had high comorbidity burden for hypertension, diabetes, obesity and CAD / MI / CHF. Table 2 provides univariable comparison of African Americans vs. Non-African Americans across various socio-demographic and comorbidity characteristics.

**Table 2:** Univariable comparison of socio-demographic and comorbidity factors between African American and non-African American

|  | **African**<br>**American**<br>**n = 1,159** | **Non-African**<br>**American**<br>**n = 3,354** | **OR / Median**<br>**Difference (95% CI)** | **P value** |
| --- | --- | --- | --- | --- |
| Age: Mean (SD) | 48.8 (17.7) | 51.2 (19.2) | 0.99 (0.99 – 0.99) | < 0.001 |
| **Age Category (%)** |  |  |  |  |
| Up to 35 | 23.6 | 23.3 | Reference |  |
| 36 – 50 | 30.6 | 26.3 | 1.25 (1.04 – 1.50) | 0.02 |
| 51 – 75 | 37.6 | 35.3 | 1.14 (0.96 – 1.36) | 0.14 |
| > 75 | 8.2 | 13.1 | 0.67 (0.52 – 0.87) | 0.003 |
| **Females** | 66.4 | 60.6 | 1.28 (1.12 – 1.48) | <0.001 |
| **Median (IQR) Zip**<br>**Income** | 60,765<br>(46,801 – 76,163) | 75,793<br>(57,252 – 102,008) | -15,028<br>(-1,667, -12,388) | < 0.001 |
| **Median Zip Income Pentiles (%)** – Pentiles of increasing Income |  |  |  |  |
| Category I | 31.8 | 15.7 | Reference |  |
| Category II | 22.4 | 17.2 | 0.64 (0.53 – 0.79) | < 0.001 |
| Category III | 22.0 | 21.5 | 0.50 (0.41 – 0.61) | < 0.001 |
| Category IV | 11.7 | 22.4 | 0.26 (0.21 – 0.32) | < 0.001 |
| Category V | 12.0 | 23.2 | 0.25 (0.20 – 0.32) | < 0.001 |
| **Population Density for**<br>**Zip: Median (IQR)** | 3256.8<br>(2123.6 – 4439.7) | 2814.2<br>(1439.1 – 4260.9) | 442.6<br>(306.7 – 578.5) | < 0.001 |
| **Population Density for Zip Pentiles (%)** – Pentiles of increasing population density |  |  |  |  |
| Category I | 12.3 | 22.6 | Reference |  |
| Category II | 23.0 | 18.2 | 2.32 (1.85 – 2.93) | < 0.001 |
| Category III | 21.3 | 19.0 | 1.95 (1.54 – 2.46) | < 0.001 |
| Category IV | 22.0 | 19.0 | 3.12 (1.69 – 2.68) | < 0.001 |
| Category V | 21.5 | 20.1 | 1.97 (1.59 – 2.48) | < 0.001 |
| Hypertension | 51.7 | 39.1 | 1.69 (1.46 – 1.91) | < 0.001 |
| Diabetes | 25.0 | 20.5 | 1.29 (1.10 – 1.51) | 0.001 |
| Obesity | 9.8 | 7.4 | 1.37 (1.09 – 1.73) | 0.008 |
| CAD / MI / CHF | 19.1 | 15.5 | 1.28 (1.07 – 1.52) | 0.006 |

### Multivariable Model and Marginal Probabilities for likelihood of SARS-Co V-2 infection

The significantly higher likelihood of SARS-CoV-2 infection among African Americans (compared to White) persisted after controlling for other demographics, insurance type, median household income, population density, and comorbidities. Adjusted odds ratios (CI) for African American vs. White: 1.84 (1.49-2.27). Our fully adjusted model estimated that Asians (vs. White) were also at a significantly higher risk of SARS-CoV-2 infection (aOR, CI: 1.46, 1.09–1.95). Furthermore, we also observed a statistically significant association between SARS-CoV- 2 infection and Hispanic ethnicity, aOR (CI): 1.70 (1.35-2.14). Higher risk of infection among males (compared to females) and higher likelihood of SARS-CoV-2 infection among elderly also remained statistically significant. Detailed output of the fully adjusted logistic regression model is presented in Table 3. The influence of African American race (vs. White) and Hispanic (vs. Non-Hispanic) ethnicity was observed uniformly across the age spectrum of 10 - 80 years. In other words, we did not observe effect modification by age for relationship between race / ethnicity and SARS-CoV-2 infection. However older age in itself remains significantly associated with higher likelihood of SARS-CoV-2 infection. Based on the marginal probabilities obtained from our fully adjusted model, the probability of SARS-CoV-2 infection in a 40-year-old African American is 18.8% whereas it is 12.9% in a 40-year-old White individual, all other adjusted variables being constant. At the age of 75 this probability is 29.0% for an African American, and 20.8% for a Caucasian. A similar relationship differential was observed for Hispanic vs. non-Hispanic. Probability of SARS-CoV-2 infection for African American vs. White, and for Hispanic vs. Non-Hispanic across age spectrum is presented in Figure 2 and Figure 3.

**Figure 2:**
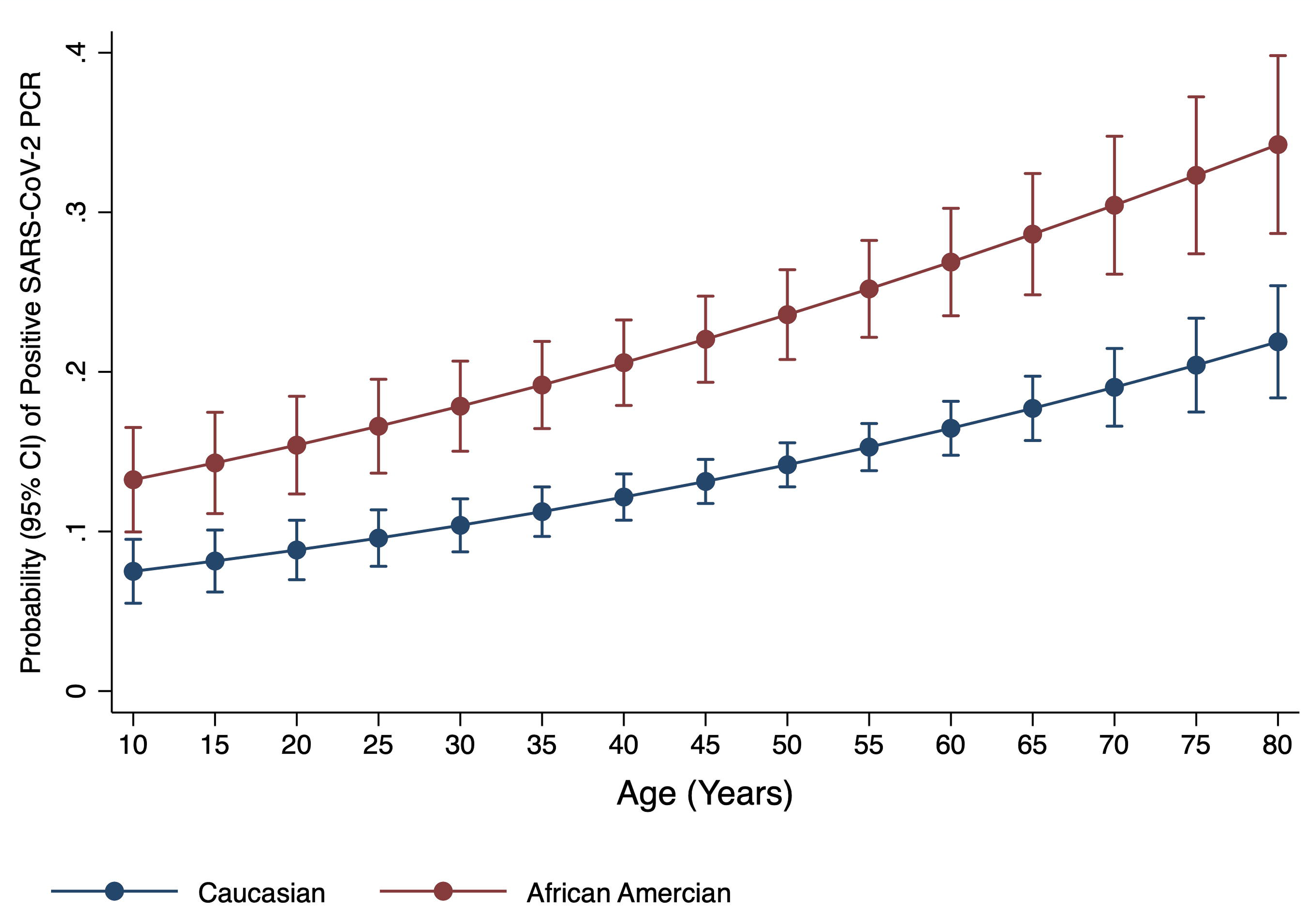
Adjusted Probability and 95% Confidence Interval of Positive SARS-CoV-2 PCR in African American vs. Caucasian by increasing age

**Figure 3:**
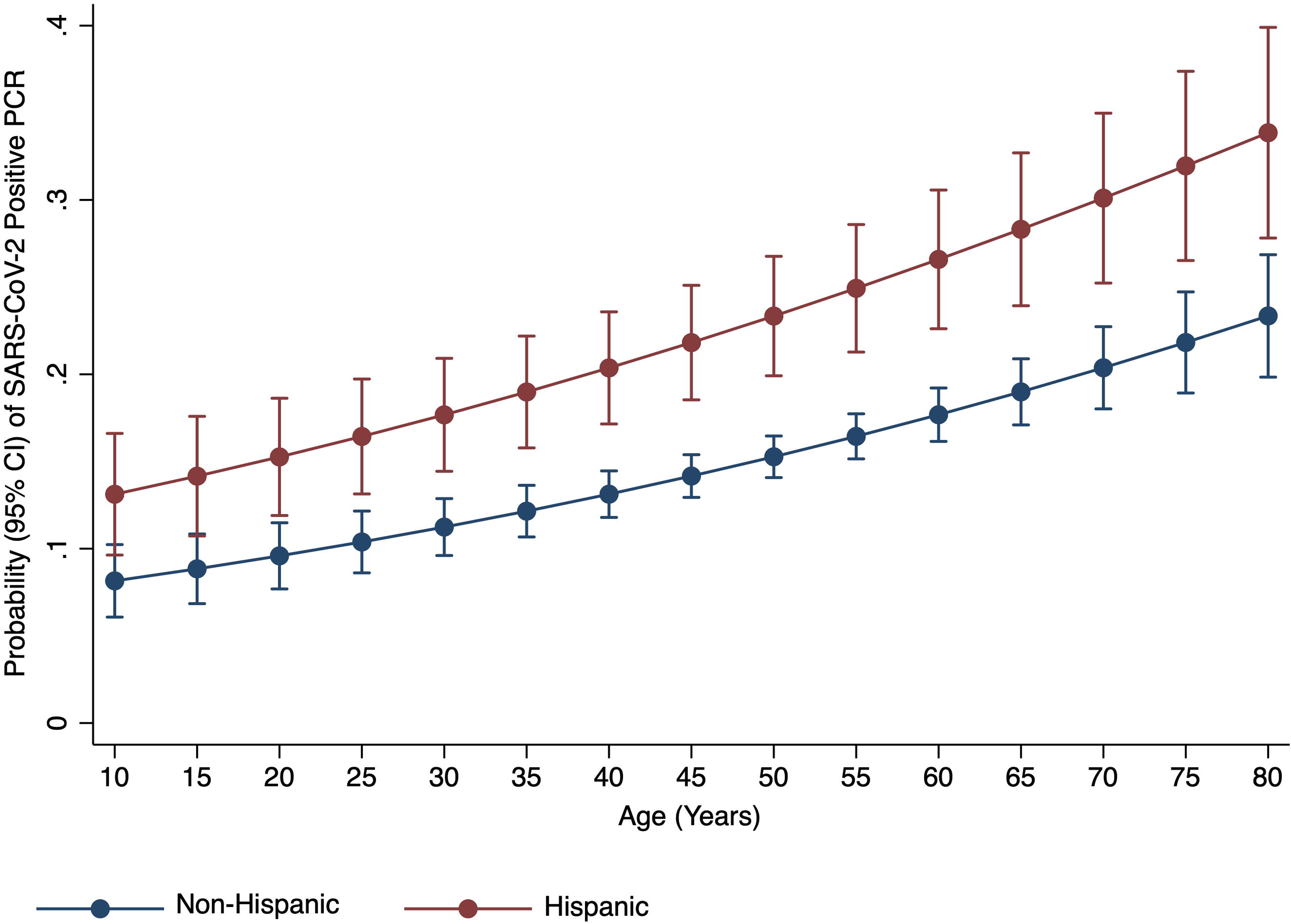
Adjusted Probability and 95% Confidence Interval of Positive SARS-CoV-2 PCR in Hispanic vs. Non-Hispanic by increasing age

**Table 3:** Adjusted Odds Ratios and 95% Confidence Intervals for socio-demographic and comorbidity factors associated with SARS-CoV-2 infection

| Covariate | Adjusted Odds Ratio | 95% Confidence Interval | P value |
| --- | --- | --- | --- |
| **Age Categories** |  |  |  |
| Up to 35 years | *Reference Category* |  |  |
| 36–50 years | 1.24 | 0.96 – 1.58 | 0.09 |
| 51–75 years | 2.02 | 1.57 – 2.61 | <0.001 |
| >75 years | 1.87 | 1.28 – 2.73 | 0.001 |
| **Male (vs. Female)** | 1.32 | 1.12 – 1.57 | 0.001 |
| **Race Categories (Non-Hispanic)** |  |  |  |
| White | *Reference Category* |  |  |
| African American | 1.84 | 1.49 – 2.27 | <0.001 |
| Asian | 1.46 | 1.09 – 1.95 | 0.01 |
| More Than One Race / Other / Unknown | 0.56 | 0.34 – 0.90 | 0.02 |
| **Hispanic (vs. Non-Hispanic)** | 1.70 | 1.35 – 2.14 | <0.001 |
| **Median Zip Household Income Categories (Pentiles of Increasing Income)** |  |  |  |
| Category I | *Reference Category* |  |  |
| Category II | 0.96 | 0.74 – 1.25 | 0.77 |
| Category III | 0.88 | 0.68 – 1.15 | 0.36 |
| Category IV | 0.88 | 0.67 – 1.17 | 0.39 |
| Category V | 0.98 | 0.74 – 1.30 | 0.91 |
| **Primary Insurance Type** |  |  |  |
| Medicare | *Reference Category* |  |  |
| Medicaid | 0.69 | 0.40 – 1.17 | 0.17 |
| Private / Employer Based | 1.18 | 0.92 – 1.51 | 0.21 |
| Healthcare Exchange | 1.58 | 0.96 – 2.60 | 0.30 |
| Self-Pay | 1.16 | 0.87 – 1.54 | 0.30 |
| Veterans Affairs | 0.22 | 0.05 – 0.93 | 0.04 |
| **Zip Population Density (Pentiles of Increasing Density)** |  |  |  |
| Category I | *Reference Category* |  |  |
| Category II | 1.08 | 0.82 – 1.44 | 0.58 |
| Category III | 1.30 | 0.99 – 1.71 | 0.06 |
| Category IV | 1.32 | 1.00 – 1.74 | 0.05 |
| Category V | 1.34 | 1.01 – 1.77 | 0.04 |
| Hypertension | 0.86 | 0.70 – 1.06 | 0.15 |
| Diabetes | 1.27 | 1.03 – 1.57 | 0.03 |
| Obesity | 0.95 | 0.70 – 1.30 | 0.75 |
| CAD / MI / CHF | 0.70 | 0.55 – 0.91 | 0.007 |

\*Hosmer and Lemeshow goodness of fit p-value: 0.63 ( $H_0$ : Model fit is correct)

### Generalized Structural Equation Modeling for Mediation by Income, Population Density and Comorbidity

Utilizing the GSEM framework, we determined the direct and indirect effects of African American race on SARS-CoV-2 infection with median income, population density and comorbidity score modeled as mediators in three separate equations. The indirect effect of African American race mediated through population density was statistically significant (p = 0.008); however, the indirect effects mediated via median income and comorbidity scores were not statistically significant (p = 0.31 and p = 0.38 respectively).

## Discussion

There is emerging evidence of race disparities in the evolving COVID-19 pandemic across the continental U.S. Most reports indicate higher case fatality among African Americans across major U.S. metropolitan areas.^9-11^ However, robust insights on the racial and ethnic differences for SARS-CoV-2 infection are limited. This is perhaps because of comparatively homogenous populations in non-U.S. regions of the world. Houston, as an exceptionally ethnically diverse population center,^14^ is well suited for an investigation of racial, ethnic, and socioeconomic gradients in COVID-19 test positivity.

Our study adds to the current literature by analyzing emerging data for individuals being tested across one of the largest healthcare systems in the Greater Houston area; we report that racial minorities (non-Hispanic African American and Asian) are approximately 50-80% more likely to test positive for SARS-CoV-2 than the non-Hispanic White population. Our data also indicate that the Hispanic population is almost 70% more likely than non-Hispanics to be susceptible to SARS-CoV-2 infection. These findings illuminate systematic racial / ethnic disparities in testing positive for SARS-CoV-2 infection. Though there are limited prior SARS-CoV-2 data, such race and ethnic disparities have previously been described for the U.S. H1N1 influenza pandemic.^15^ These data indicated that Spanish-speaking Hispanics were at a greater risk of H1N1 infection primarily attributable to lack of healthcare access. Black people were also more susceptible to complications of H1N1 infection.^15^

We explored three possible mechanisms of race disparities in our data. These included lower socio-economic status, residence in higher population dense areas, and higher level of comorbidities. We demonstrate that African American race is significantly associated with all three potential disparity pathways, and in the traditional multivariable analyses, race and ethnic disparities persisted even after controlling for these pathways. However, our mediation analyses highlighted the potential influence of residence in high population density areas as a viable pathway that at least partially explains race disparity. Pathways mediating the influence of median income and comorbidity status did not demonstrate a significant effect. We utilized population density as a marker for potential inability to maintain adequate social distancing as it has been indicated that maintaining the WHO recommended safe distance between people becomes challenging with high population densities.^16^ Furthermore, overall effects of population density and disease spread has been previously described in literature.^17,18^ In addition to lack of social distancing, higher population density may also be associated with several other behavioral and socio-demographic attributes that may predispose to both viral spread and increased susceptibility. For example, there are reports linking obesity, lack of physical activity, and higher mortality with residence in densely populated neighborhoods.^19,20^

As reported, our data also corroborate that older populations may be more susceptible to SARS-CoV-2 infection;^8^ however, younger populations still have cause for concern, as nearly 1 in 4 of the infected cases in our sample were between 36-50 years. Finally, our data demonstrate that males may be approximately 30% more likely to test positive for the SARS-CoV-2 infection. Potential sex differences in infectivity to SARS-CoV-2 and intersectionality with racial and ethnic socioeconomic factors need to be explored further in future analyses. Additional policy-oriented research should prioritize study on the intersectionality of these vulnerable economic statuses and racial disparities in COVID infection indicated by the present study.

Findings of our study need to be interpreted in the light of certain limitations. First, our data are from a single center and may not be generalizable to the wider U.S. population. These findings need to be replicated in larger data sets across other large heterogenous U.S. metropolitans. However, the Houston metropolitan area is one of the most diverse and representative in the U.S.,^14^ and our healthcare system is one of the largest systems providing care to COVID-19 patients in the Greater Houston area. Our sample was composed of 26% Black, 19% Hispanic, and 62% female population. Second, we did not have information on certain demographic covariates such as education. Educational status has been linked to healthcare awareness and may be important to adjust for in analyses of potential disparities. However, we obtained and adjusted for zip code income data from the U.S. Census, as income has previously been shown to have strong correlation with educational attainment.^21^ Third, since testing was based on suspicion of infection and may have been influenced by factors such as access to care, the potential for selection bias cannot be ruled out. Finally, we did not have detailed information on comorbidities and their management in the study population. However, we did control for major comorbidities which are being reported as associated with COVID-19 outcomes.^22^

## Conclusions

The strong association between racial and ethnic minorities and SARS-CoV-2 infection demonstrated in our data, even after adjustment for other important socio-demographic and comorbidity factors, highlight a potential catastrophe of inequality within the existential crisis of a global pandemic. Our data, representing a large heterogeneous U.S. metropolitan area, also provide preliminary evidence into the potential pathways for this disparity. It is highly likely that higher comorbidity burden and detrimental effects of adverse social determinants, including those that may not adequately permit safe practices of social distancing, mediate higher SARS-CoV-2 infectivity among racial and ethnic minorities.

As the pandemic continues to spread and evolve across the continental U.S., emerging data on association between SARS-CoV-2 infection and various socio-demographic factors will continue to enhance our understanding of targeted risks related to SARS-CoV-2 infection, and such data would enable us to comprehend healthcare services and access factors related to development and outcomes of COVID-19 among minority populations.

## Data Availability

Data requests can be made to the corresponding author and will be evaluated by the appropriate institutional committees on a case by case basis.

## Acknowledgment

The authors thank Jacob M. Kolman, Senior Scientific Writer of the Houston Methodist Center for Outcomes Research, for reviewing the language and format of the manuscript.

## IRB Approval

This work was carried out under an approved protocol for the Houston Methodist COVID-19 Surveillance and Outcomes Registry (HM CURATOR) by the Houston Methodist Research Institute Institutional Review Board (HMRI IRB).

## Contribution Statement

FV: design, data analysis and interpretation, drafting the manuscript, critical revision for important intellectual content, final approval

JCN: data acquisition, data analysis, drafting the manuscript, final approval

OK: data acquisition, data analysis, drafting the manuscript, final approval

SLJ: data acquisition, data interpretation, critical revision for important intellectual content, final approval

FNM: critical revision for important intellectual content, final approval

HDS: critical revision for important intellectual content, final approval

RAP: critical revision for important intellectual content, final approval

JDA: critical revision for important intellectual content, final approval

BK: critical revision for important intellectual content, final approval

KN: design, interpretation of data, critical revision for important intellectual content, final approval

## Date sharing statement

All requests for de-identified data should be made to the corresponding author. All reasonable requests will be evaluated by the CURATOR Data Governance and Sharing Committee comprising of FV, SLJ, BK and KN in the light of institutional policies and guidelines.

